# Effects of improved water, sanitation, handwashing and nutrition on early childhood IgG immune repertoire development against *Shigella* and enteroinvasive *Escherichia coli* (EIEC)

**DOI:** 10.1101/2025.11.21.25340591

**Authors:** Nikolina Walas, Everlyn Kamau, Minlu Zhang, Kathy Kamath, Jack Reifert, John Shon, Shahjahan Ali, Md. Ziaur Rahman, Abul K. Shoab, Syeda L. Famida, Salma Akther, Md. Saheen Hossen, Palash Mutsuddi, Mahbubur Rahman, Andrew N. Mertens, Richelle Charles, Daniel Leung, Stephen Luby, Audrie Lin, Benjamin F. Arnold

## Abstract

We studied effects of a combined water, sanitation, handwashing, and nutritional supplementation (WSH+N) intervention on *Shigella/*EIEC IgG immune development among Bangladeshi children enrolled in a cluster randomized trial. We used a bacterial display assay to measure IgG-specific binding to *Shigella*/EIEC Ipa proteins (IpaA, IpaB, IpaC, IpaD, IpaH) from a substudy of 120 children (60 intervention, 60 control) at median ages 3-, 14-, and 28-months. We found that the WSH+N intervention did not impact IgG seroprevalence (56% vs 53%, P=0.6) or IgG epitope repertoire development (P=0.15-0.97), but there were distinct age-based patterns of IgG linear epitopes and motifs charting the development of the immune repertoire. Samples from before age 6-months, reflecting predominantly maternal IgG, had higher epitope breadth and magnitude compared with samples from older ages. A modeling analysis of maternal-child IgG dynamics suggests peak susceptibility occurs between 6 and 9 months of age. With this methodology, we find that intensive, household-based WASH interventions were insufficient to reduce early childhood infection to *Shigella*/EIEC. Our results motivate complementary interventions, such as vaccines, to prevent *Shigella*/EIEC infection in the first two years of life.

## Introduction

*Shigella* and enteroinvasive *Escherichia coli* (EIEC) are closely related bacterial pathogens, estimated to cause 111 million cases and 63,000 deaths annually in children under the age of five (Anderson et al., 2019; Libby et al., 2023; Sriguha et al., 2025). *Shigella/*EIEC are symptomatically and biochemically difficult to distinguish and are often grouped together when evaluating diarrheal causes (Beld & Reubsaet, 2012; Ud-Din & Wahid, 2014). Antibiotics are the only treatment option due to no available vaccine. Antibiotic use creates selective pressure for antimicrobial resistance, which is largely driven through horizontal gene transfer in *Shigella/*EIEC species (Ahmed et al., 2023; Baker et al., 2018). In Bangladesh, the prevalence of resistance to first-choice antibiotics is estimated to be as high as 62%, with an overall prevalence of multi drug resistant *Shigella* at 33% (Azmi et al., 2014; Puzari et al., 2018) (Ahmed et al., 2023), (Raso et al., 2023). The burden associated with *Shigella/*EIEC and development of antimicrobial resistance motivate preventive interventions to reduce infections in high transmission settings.

*Shigella/*EIEC is primarily transmitted fecal-orally and due to its low infectious dose (10-100 bacteria), outbreaks can spread quickly from person to person or through contaminated water in settings with poor sanitation (Shad & Shad, 2021; Zaidi & Estrada-García, 2014). Household level behavioral interventions such as handwashing, clean drinking water, and human waste sanitation have been found to reduce diarrheal incidence (Aiello et al., 2008; Curtis et al., 2000), though these results have not been consistent across populations in low- and middle-income settings (Humphrey et al., 2019; Null et al., 2018). In the WASH Benefits Bangladesh trial, WSH+N interventions did not significantly reduce *Shigella*/EIEC infection at 14-months measured by PCR (Grembi et al., 2023). This null finding may be due to seasonal variability in enteric pathogen prevalence and delayed sample collection between comparison groups due to political unrest, though similar nonsignificant results were observed in other study populations (Muriithi et al., 2024; Rogawski McQuade et al., 2020). Single measures of pathogen presence in stool likely underestimate the cumulative incidence of actual infections. Immunoglobulin G (IgG) antibodies to *Shigella*/EIEC measured in blood are long-lived and provide a cumulative measure of previous infection that could complement PCR-based measures from stool (Arnold et al., 2018; Bernshtein et al., 2024).

Here, we aimed to assess the impact of the combined nutritional and water, sanitation and hygiene (WSH+N) intervention from the WASH Benefits Bangladesh trial on *Shigella*/EIEC IgG using a randomized bacterial display assay and the Serum Epitope Repertoire Analysis (SERA) platform. We hypothesized that improved water quality, handwashing and sanitation interventions would reduce exposure to *Shigella/*EIEC in early childhood and therefore result in lower IgG immune responses. We characterize the age-specific repertoire of linear epitopes and conformational motifs of IgG antibodies specific for *Shigella* Ipa proteins (IpaA, IpaB, IpaC, IpaD, IpaH) in children sampled longitudinally at 3-months, 14-months and 28-months. We model the waning of maternally derived antibodies and the subsequent rise in seroprevalence due to natural infection. Finally, we estimate the window of infection susceptibility for the study population. Our results provide new estimates of the impact of WASH interventions on *Shigella/*EIEC immune development and could help inform future vaccine timing.

## Methods

### Ethics

Written, informed consent from participants was provided before enrollment and before specimen collection. The Ethical Review Committees at the International Centre for Diarrheal Disease Research, Bangladesh (PR-11063), the Committee for the Protection of Human Subjects at University of California, Berkeley (2011-09-3652), the Institutional Review Board at Stanford University (25863) and the University of California, San Francisco (22-36722) approved the protocol for the original study (Clinical Trial Registration NCT01590095).

### Study design and sample selection

The WASH Benefits Bangladesh trial was a cluster-randomized trial to investigate the independent and combined effects of improved child nutritional and household WASH on children’s linear growth and diarrhea. Details for study design and enrollment are described elsewhere (Arnold et al., 2013). In brief, geographically matched clusters were randomized to one of six intervention arms (improved water; improved sanitation; improved handwashing; improved nutrition; improved water, sanitation and handwashing; or improved water, sanitation, handwashing and nutrition) or the control arm. Within the larger trial, a substudy focused on environmental enteric dysfunction enrolled 217 clusters from four arms (control; improved water, sanitation and handwashing; improved water, sanitation and handwashing and nutrition; and improved nutrition) of the trial and collected venous blood samples from the birth cohort via household visits by field staff at three follow up visits (median child ages 3-months, 14-months and 28-months) and plasma samples were stored according to previously published methods (Lin et al., 2020).

We selected a sample of 120 children for the immune repertoire substudy (**Table S1**). We restricted our analysis to children that were enrolled in the control group or the combined nutritional and water, sanitation and hygiene (WSH+N) intervention group (n=900) and had a blood sample collected at all three visits. We excluded children that had their first measurement (ages 1-6 months) between August and November of 2013 to improve comparability between arms in visit timing with respect to calendar date and season, as measurement in control villages was paused during late 2013 due to political instability in Bangladesh. We also restricted the sample to children who were aged between 28 and 200 days at visit 1 and 365 and 548 days at visit 2 to improve balance between the two groups with respect to age and measurement dates. Among 147 children who met inclusion and exclusion criteria, we selected a random sample of 120 children (60 children per group, 360 total samples) for the analysis. Overall sample size was determined primarily based on logistical and cost feasibility for the immune repertoire assay testing. We determined a sample size of 60 children per group would have 80% power to detect an absolute reduction of 25% seroprevalence with a two-sided alpha of 0.05 and an assumed 50% seroprevalence using a standard equation for comparison of two proportions.

### Serum Epitope Repertoire Analysis (SERA) Platform

The peptide display library was prepared and used for selection according to detailed methods previously published (Kamath et al., 2020; Reifert et al., 2021). Briefly, the *E. coli* peptide display library was induced for peptide expression, resuspended in a 15% glycerol/PBS solution (PBSG) and aliquoted in deep well 96-well plates at 10-fold the library diversity in each well (8×10^10^ cells per well). Plates were frozen and stored at – 80 °C for future use. Selection was performed per published protocol using a 1:25 dilution of each sample, capture of antibody bound bacteria using protein A/G Sera-Mag SpeedBeads (GE Life Sciences, 17152104010350) (6.25% the beads’ stock concentration), and growth of the selected bacteria overnight with shaking (300 rpm) at 37 °C.

Plasmids were isolated for the selected bacteria and PCR was used to amplify the region encoding the random peptides expressed on the cell surface during selection and to incorporate adaptors and indices necessary for pooling samples for NGS sequencing. Details have been previously published (Kamath et al., 2020; Reifert et al., 2021). Purified amplicons with indices and adaptors were pooled, prepared for sequencing and sequenced according to specifications of the Illumina NextSeq 500 using a High Output v2, 75 cycle kit (Illumina, FC-404-2005).

### Bioinformatics

Once samples were processed through the SERA assay and sequenced, they were scored against all previously developed and validated epitope motifs reflective of antibody responses to different exposures. Previously published works have described in detail methods for motif discovery using an algorithm (IMUNE) to create and validate exposure panels (Kamath et al., 2020; Pantazes et al., 2016; Reifert et al., 2021). Briefly, groups of epitope motifs (panels) that demonstrate high sensitivity, specificity, and breadth of coverage in a discovery disease and control set of samples are selected and compiled together. A control group is used to generate z-scores for every motif and every sample screened through the assay. Individual motif z-scores are summed for each unique epitope to generate a composite score for the exposure for which that motif panel represents. The resulting composite score (SERA score) is used to determine a cutoff for positivity based on predicate tested disease and controls. A new set of validation samples are then tested and scored to determine performance metrics (sensitivity and specificity) of the motif panel. All samples from this study were scored against motif panels developed for the detection of the listed exposures. Heatmaps of z-scores for each individual motif against every sample are provided in supplemental data to compare relative reactivities across sample sets for each antibody reactive epitope.

A panel of epitope motifs was discovered for exposure to Shigella using the same methods described above with the exception that predicate tested controls were not utilized. Instead, databased sample data originating from the US and provided as part of a cohort of Blood Centers of America (BCA) healthy controls were used in comparison to samples provided for different infectious diseases prevalent outside of the US. A selection of the samples for those born outside of the US (ex-US)were compared to BCA samples to discover motifs that aligned to known shigella antigens (primarily invasion plasmid antigens – IpaB, IpaC, and IpaD) and developed into a panel of epitope motifs as described previously (Kamath et al., 2020). Because no predicate tested validation samples were available, additional ex-US samples were used to estimate a composite score cutoff of 50 for the panel (exposure panel is preliminary).

To identify linear epitopes, we investigated the Ipa proteome using protein-based immunome wide association studies (PIWAS) (Haynes et al., 2021). For each sample, approximately 750,000 to 3.6 million unique 12-mer sequences were obtained from the SERA assay, and the sequences were decomposed into constituent 5- and 6-mers. An enrichment score for each 5- or 6-mer was calculated by dividing the number of unique 12-mers containing the kmer by the number of expected 12-mers for the sample, based on the amino acid proportions in the sample. For each sample of the study, 5- and 6-mers were tiled against protein sequences for *Shigella sonnei* Ipa proteins (**Table S2**), and a tiling enrichment score, sometimes referred to as IWAS score, was calculated at each amino acid position of the protein based on the corresponding 5- or 6-mer enrichment scores, with a smoothing window of five 5-mers and five 6-mers. We then log-transformed these enrichment scores to Z-scores and normalized to BCA health population controls as described in (Pantazes et al., 2016). The normalized z-scores are hereafter referred to as IgG enrichment values.

### Defining epitopes, seroreactive regions and summary measures

We defined epitope hits as eight or more consecutive amino acids with an enrichment tiling score above the 90^th^ percentile enrichment score for that protein. Areas of high reactivity per protein, referred to as seroreactive regions, were defined as amino acid regions that contained epitope hits in at least five unique samples. A sample was defined as being seropositive to a protein if one or more epitope hits were identified. Seropositivity to the pathogen was defined as having at least two epitope hits to any of the Ipa proteins included in the panel. Quantitative summary measures were defined as the count of epitope hits in seroreactive regions and the mean of enrichment values in seroreactive regions over the entire protein, akin to the outlier sum score proposed by Tibshirani and Hastie (Tibshirani & Hastie, 2007) for analyses of genomic expression data. To account for proteins with varying amino acid length, the pathogen level summary of the mean enrichment in seroreactive regions was taken across the amino acids in seroreactive regions for all proteins in the panel. Total epitope hits in a sample were calculated by summing over all Ipa proteins in the panel.

### Statistical analysis

Participants were analyzed according to their original randomized group (intent to treat). Primary between-group comparisons combined results from visits 2 and 3 at median ages 14-months and 28-months, excluding earlier samples to avoid potential contributions of maternal IgG. We used linear-binomial regression models to estimate the difference in IgG seroprevalence between treatment groups, accounting for repeated outcome measurements using robust standard errors at the cluster level. The same approach was used to estimate the difference in seroconversion per child-year, but we assumed all children were at risk of seroconversion at visit 2 and only those seronegative at visit 2 were at risk for seroconversion between visits 2 and 3. We used linear regression models to estimate the difference of quantitative summary measures, mean IgG enrichment, maximum IgG enrichment, and the SERA score between groups, while accounting for repeated measures using robust standard errors. Differences in epitope hits between the groups was estimated using a permutation test based on the Mann-Whitney U test statistic, with treatment permuted at the cluster level (10,006 permutations).

We used a modified catalytic model framework to account for passive maternal IgG transfer and IgG developed from natural infection and estimate age-based *Shigella*/EIEC seroprevalence as described in depth elsewhere (Kamau et al., 2025). Briefly, the model assumes a constant force of infection over time and age, defined as the rate at which susceptible individuals become infected (parameter λ), and a constant rate of decay of maternal antibodies (parameter γ) using a closed system of differential equations outlined below,

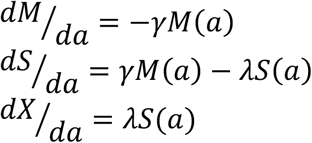

where λ > 0; γ > 0;

The system solution has the following solutions:

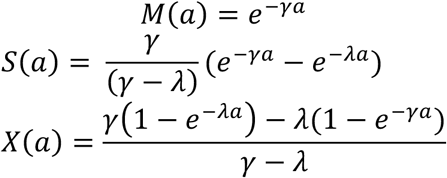

Where,

M(a) is the proportion of children of age *a* losing maternal immunity at a constant rate γ, S(a) is the proportion of children susceptible among those born with maternal immunity, and X(a) describes the proportion of children seropositive from natural infection at a constant rate λ.

The models were implemented in a Bayesian framework using Rstan (Stan Development Team, n.d.). We used a strongly informative prior for γ, specified as a normal distribution with a mean of 0.5 and a variance of 0.001, and a moderately informative prior for λ, specified as a normal distribution with a mean of 0.1. Parameter posterior distributions were estimated using four chains each with 3000 iterations and 900 burn-in. Monte Carlo Markov chain convergence was assessed by Gelman-Rubin R-hat diagnostic with a threshold of 1.1. A threshold of over 400 was used for effective sample size. Means of parameter estimates and 95% credible intervals (2.5% and 97.5% percentiles of posterior distribution) are presented. Model fit is presented in supplemental figure 2.

## Results

### Participant characteristics and approach to epitope mapping

Our analyses are based on longitudinal serum samples collected from a subsample of 120 children that were selected from the control and the WSH+N intervention group of the WASH Benefits Bangladesh trial. We developed a pipeline to identify IgG linear epitopes against five *Shigella*/EIEC Ipa proteins (IpaA, IpaB, IpaC, IpaD and IpaH) using amino acid level enrichment data from a randomized bacterial display assay (**Figure 1, Table S1**). Epitope hits were defined as eight or more consecutive amino acids with an enrichment tiling score above the 90^th^ percentile enrichment score for that protein. Seroreactive regions, were defined as amino acid regions that contained epitope hits in at least five unique samples. Seropositivity to the pathogen was defined as having at least two epitope hits in seroreactive regions to any of the Ipa proteins included in the panel. Sensitivity analyses suggest similar age-dependent patterns were slightly elevated or blunted depending on the chosen threshold for defining seroreactive regions and pathogen level prevalence. We chose the current threshold as it was an intermediate for our reported estimate and clearly maintained the observed age dependent pattern in this population (**Figure S1**). In parallel, we identified epitope motifs, motif families and summarized a composite score, referred to here as the SERA score, determined by the IMUNE algorithm as detailed in our methods.

**Figure 1.**
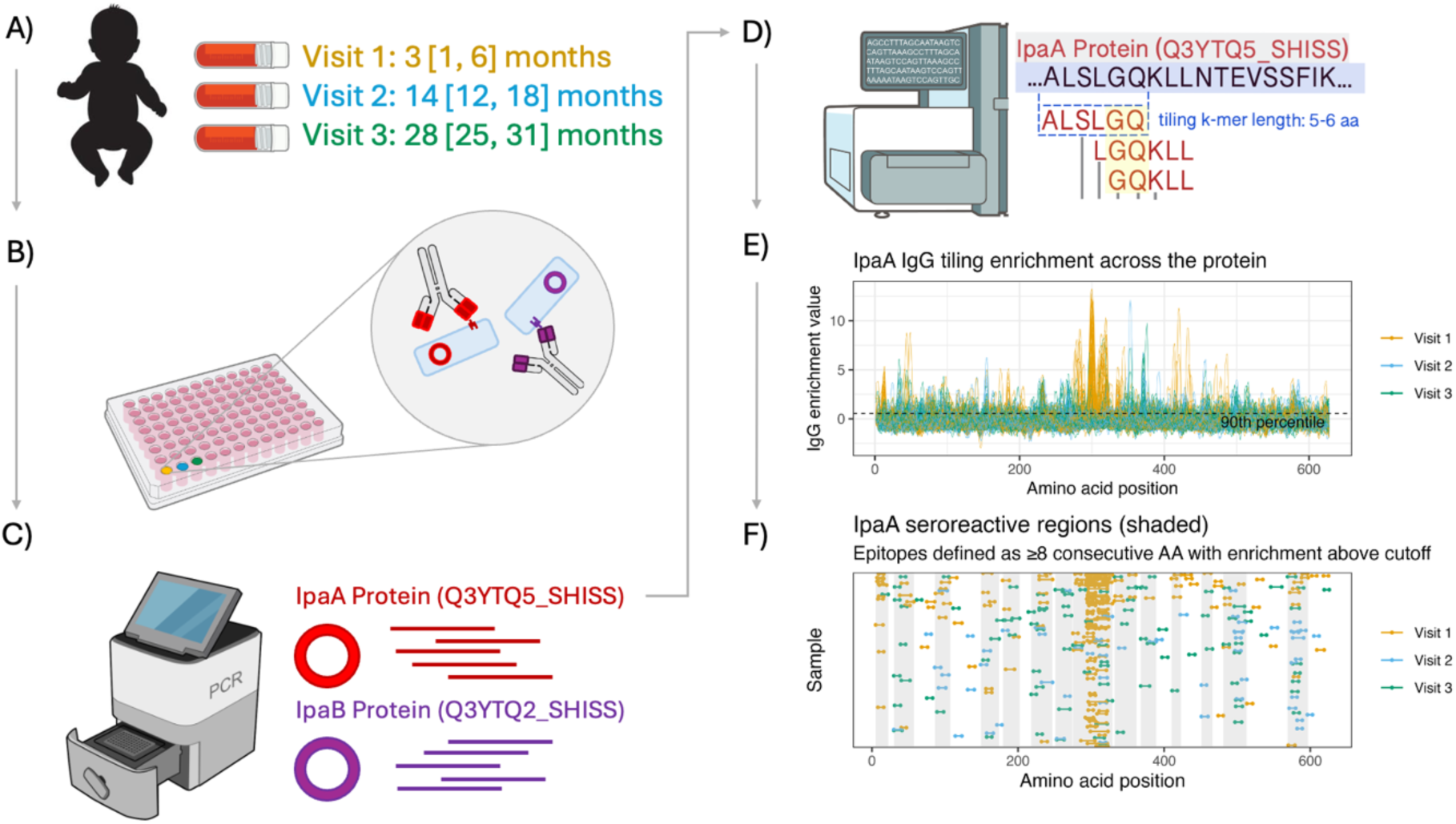
Overview of linear IgG epitope identification. (A) Three hundred and sixty serum samples originating from a sample of 120 children were analyzed using a randomized bacterial display assay. (B&C) Plasmids expressing protein sequences that bound serum antibodies were purified and sequenced using PCR. (D) An IgG tiling enrichment score was calculated for each amino acid position of corresponding *Shigella*/EIEC Ipa proteins (IpaA, IpaB, IpaC, IpaD, IpaH) using 5-mer and 6-mer log-enrichment with a window size of 5 for smoothing and normalized to healthy controls (E) Epitope hits were defined as eight or more consecutive amino acids with an IWAS score above the 90^th^ percentile for that protein across all 360 serum samples. (F) Seroreactive regions were defined as amino acid regions with an overlap of 5 or more epitope hits from unique samples (1.5% of samples).

Participant characteristics were generally well balanced between intervention and control groups in the substudy (**Table 1**). Compared with intervention group, the control group had more females (57% versus 52%) and mothers had more years of education (6.2 years versus 4.9 years). Overall, household socioeconomic status and WASH characteristics were generally similar between the groups. Measurement age was similar between groups across the three longitudinal visits, with control group children approximately one month older on average at visits 1 and 2 (**Table 1**).

**Table 1.**
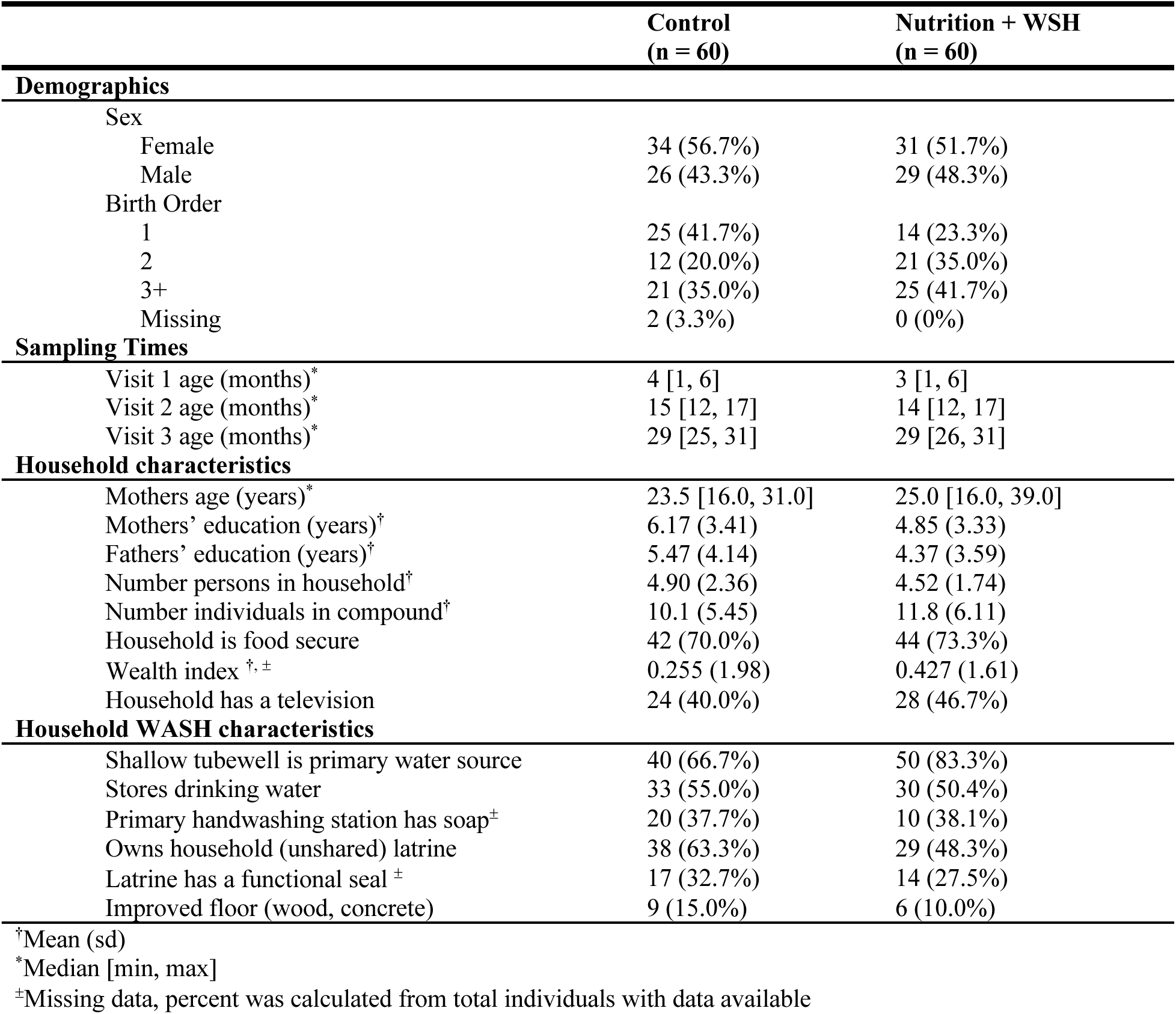
Baseline characteristics of control and intervention groups enrolled in the substudy.

#### 4.4.2 Intervention effects on Shigella/EIEC IgG

We assumed that samples collected at median age 3-months predominantly reflected maternal IgG acquired in utero (Allansmith et al., 1968; Zinkernagel, 2001). Estimates of intervention effects thus focused on sera collected at ages 14- and 28-months, which we assume measure IgG responses following incident infections. Seroprevalence in the control group declined from 85% at 3-months to 51% at 14-months and 28-months (Figure 2A), likely reflecting a combination of maternal IgG decay and newly produced IgG as a result of incident infections, which will be examined in detail below. Age-dependent patterns were nearly identical in the intervention group, and IgG seroprevalence and seroconversion rates estimated across ages 14- and 28-months did not differ between children in intervention and control households (**Table S3**).

**Figure 2.**
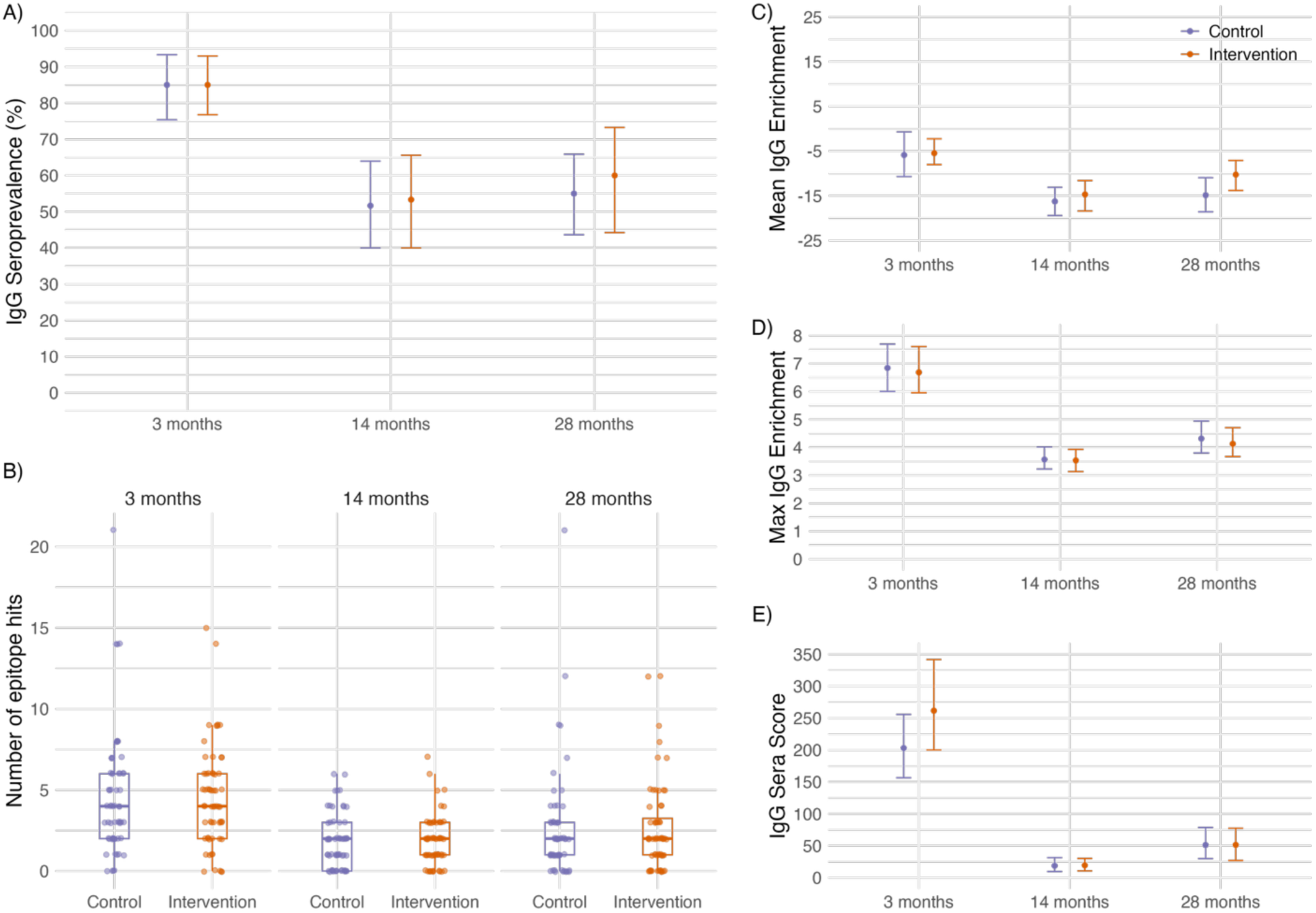
IgG responses to *Shigella*/EIEC Ipa proteins and motifs by intervention group and age. Seroprevalence, epitope counts, and enrichment measures were aggregated across IpaA, IpaB, IpaC, IpaD, and IpaH proteins. Samples were analyzed from 3 longitudinal visits at median age at 3 [1, 6] months, 14 [12, 17] months, and 28 [25, 31] months (N=60 children per group, 360 total samples). (A) IgG seroprevalence by intervention group and age determined as ≥2 epitope hits across Ipa proteins. (B) Number of epitope hits across Ipa proteins, by intervention group and age. Box plots summarize the median and interquartile range IQR, with whiskers representing 1.5 times the IQR. Points represent individual samples (N=60 per group and age). (C) Mean IgG enrichment score, calculated as the average IgG enrichment within seroreactive epitope regions of the Ipa proteins. (D) Max enrichment score, calculated as the maximum IgG enrichment within seroreactive epitope regions of the Ipa proteins. (E) Mean SERA score, calculated as the composite score for epitope motifs identified specific to *Shigella*/EIEC through the IMUNE algorithm. There were no statistically significant differences between intervention groups overall or by age across all IgG summary measures (P>0.05). In panels A, C, D, and E, points represent means and bars represent 95% confidence intervals estimated through bootstrapping, accounting for repeated measures.

Intervention and control groups were also very similar with respect to quantitative measures of immune response, including the number of epitope hits, mean IgG enrichment, maximum IgG enrichment, or IgG SERA score (Figure 2, **Table S3 and Table S4**).

### Age-based patterns of Shigella/EIEC IgG epitope repertoire

Distinct amino acid regions with enrichment for linear epitopes were observed for each of the three ages. Generally, the magnitude of IgG enrichment was higher in 3-month samples, assumed to be the maternally transferred IgG, compared to 14-month and 28-month samples. (Figure 3A). Notably, IgG enrichment from amino acid position 290 to 325 in IpaA was 4 times higher in 3-month samples compared to 14-months and did not increase at 28-month samples. The largest epitope motif family 1, corresponded to this amino acid region (Figure 4). Areas of elevated amino acid enrichment were less distinguished by age for IpaB, IpaD, and IpaH3, though still present. Notably, epitope motifs identified through the IMUNE algorithm were present in all Ipa proteins except IpaH3 (**Table S5**), with motif families, defined as epitope motifs with overlapping amino acid positions, detected in IpaA, IpaB, and IpaC (Figure 4). The proportion of samples positive for an epitope motif was higher at 3-months compared to 14-months or 28-months for all 32 epitope motifs detected (**Table S5**).

**Figure 3.**
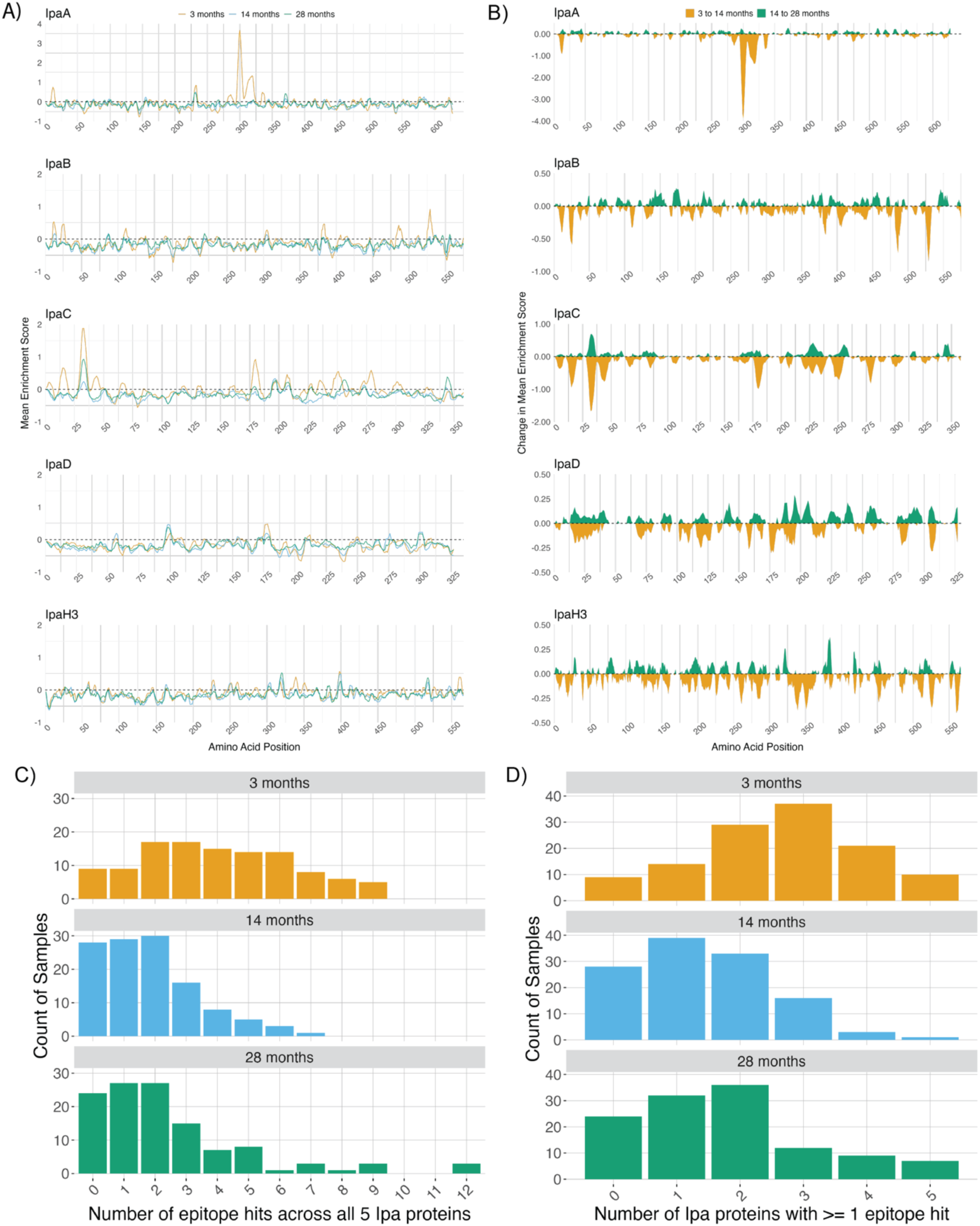
Amino acid enrichment and linear IgG epitope breadth across three longitudinal visits with children’s median age at 3 [1, 6] months, 14 [12, 17] months, and 28 [25, 31] months. (A) Measures from the first visit at age 3-months reflect maternal antibodies and show higher enrichment relative to ages 14 [12, 17] months, and 28 [25, 31] months. (B) The mean change in amino acid enrichment score was calculated by averaging the enrichment score at each amino acid region for all 3 age groups and then subtracting the calculated mean enrichment score from the preceding visit’s mean enrichment score. (C) Breadth of Ipa-specific IgG immune response as summarized by number of epitope hits is higher for 3-month samples. Six 3-month samples (epitope counts: 14 (4) 15 (1), and 21 (1) and one 28-month sample (epitope count: 21) were excluded to accommodate data visualization. (D) Breadth of Ipa-specific IgG responses as defined as the number of Ipa proteins with an epitope hit is also higher in 3-month samples.

**Figure 4:**
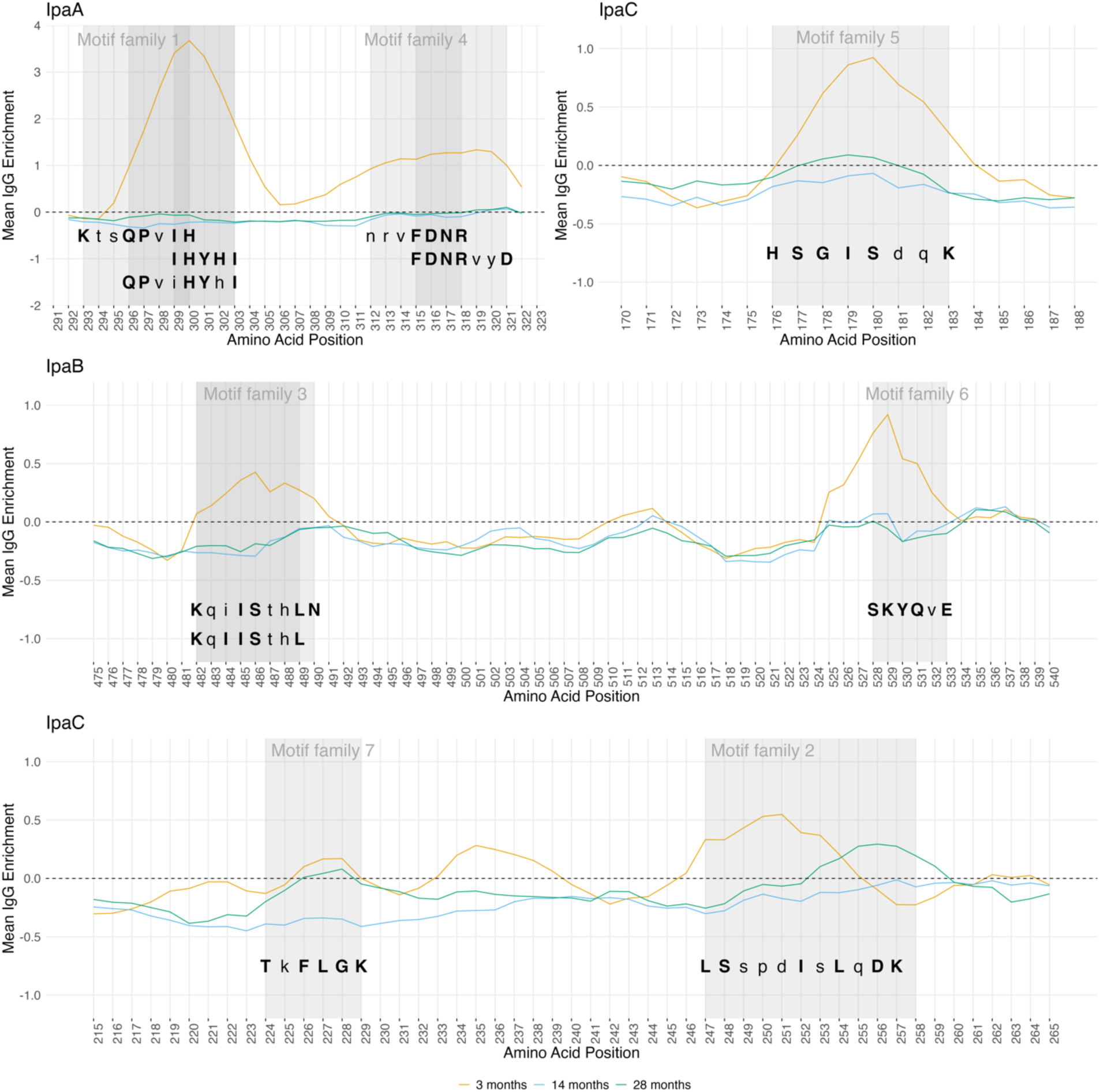
Corresponding amino acid sequences of epitope motif families. Motif sequences were identified using the IMUNE algorithm that compiled *Shigella/*EIEC epitope motifs based on a set of healthy US controls compared to non-US samples. Motif families were defined as identified epitope motifs with overlapping amino acid regions. Motif families were identified in IpaA, IpaB, and IpaC proteins. Grey shading represent the amino acid range of epitopes within each motif family. Capitalized and bolded letters of sequences represent the amino acids of the epitope motifs. Trace lines represent mean IgG enrichment tiling values per amino acid position as determined through the SERA PIWAS tiling method.

IgG epitope breadth, as defined as the count of epitope hits and the count of unique Ipa proteins with an epitope hit was higher at 3-months compared to the other timepoints (Figure 3C-D). The count of linear epitope hits per sample at 14-months and 28-months were right-skewed, with most samples having only 0, 1, or 2 epitope hits, whereas at 3-months the distribution of linear epitope hits per child were more uniformly spread over a higher count range. This pattern was also observed for the count of unique Ipa proteins with an epitope hit.

### Inferred Shigella/EIEC immunity dynamics and age-based susceptibility

We used a modified catalytic model framework to estimate *Shigella*/EIEC infection rate and maternal antibody waning rate and model age-specific seroprevalence (Figure 5). The model estimated IgG seroprevalence steadily declined from 100% seroprevalence to 44% between 6 and 12-months, corresponding with the steady loss of maternal IgG by 12 months. The estimated IgG seroprevalence attributable to maternal IgG approached 0 by 18 months with a handoff to IgG from natural infection between 6 and 9-months. Taken together, the estimated proportion of individuals seronegative, or susceptible to infection, peaked at 44% between ages 6 and 9-months.

**Figure 5.**
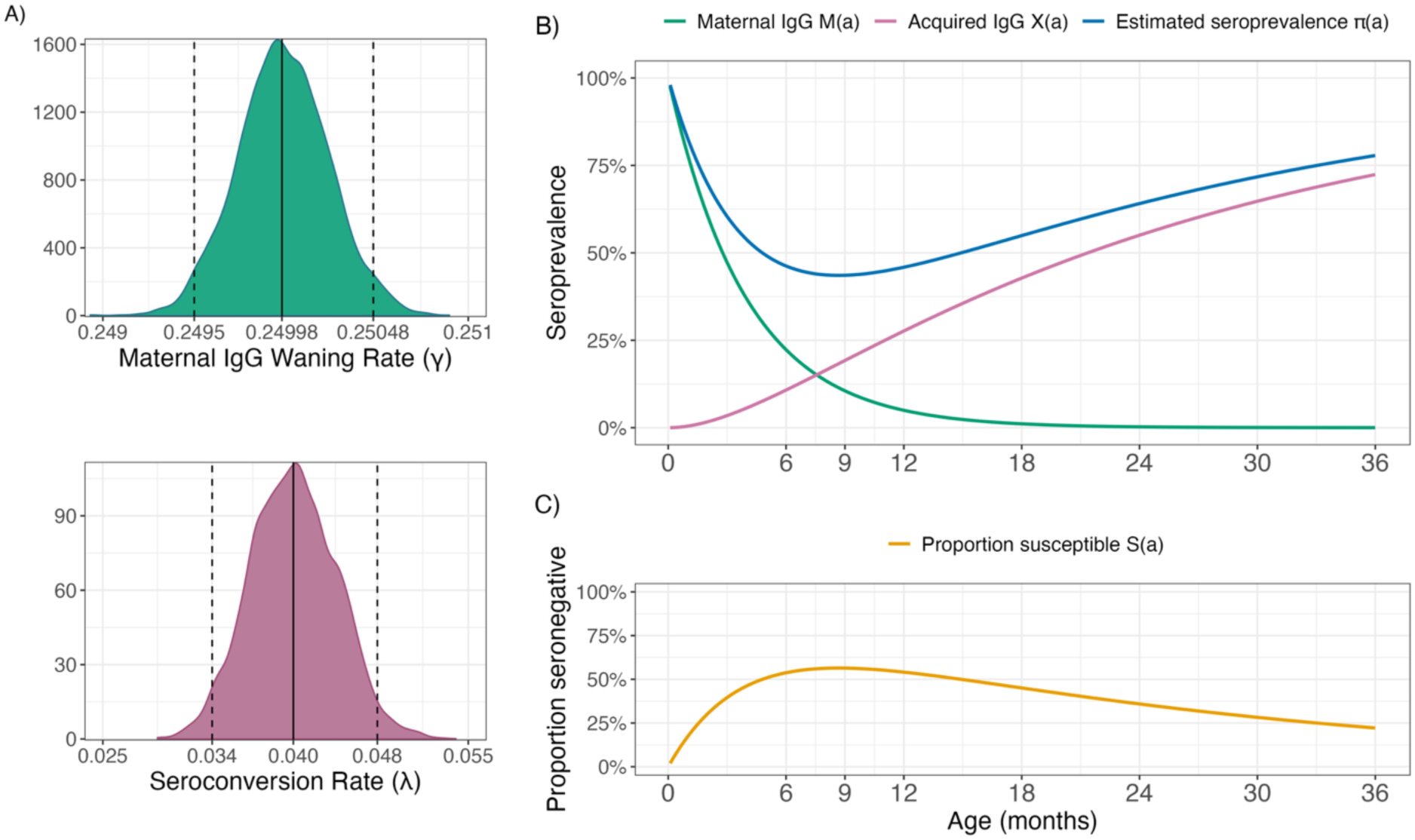
Maternal and child IgG dynamics inferred from age-dependent seroprevalence. A modified catalytic model was used to estimate the rate at which individuals become infected, the seroconversion rate (λ), and the rate at which maternal antibodies wane (γ), under the assumption that all children were born with maternal antibodies. See methods for more specifics on model building and mathematical derivations. (A) Posterior distributions of model parameters, including the median estimates (vertical line) and 95^th^ credible intervale (dashed lines). (B) These model parameters were used to estimate population immunity projections including maternal IgG, acquired IgG, and observed seroprevalence from one to 36-months. (C) The proportion of the population susceptible to *Shigella*/EIEC, assuming IgG is a correlate of protection, was also estimated from the catalytic model.

## Discussion

There was no significant impact of the WSH+N intervention on children’s IgG seroprevalence and seroconversion. Our results suggest that intensive household WSH+N interventions do not influence a child’s immune development to *Shigella*/EIEC Ipa proteins. This is consistent with an absence of intervention effect on *Shigella*/EIEC infection measured by qPCR at age 14-months in the same study population (Grembi et al., 2023). Since IgG integrates information about cumulative *Shigella*/EIEC infections over time, this study provides a comprehensive assessment of intervention effects during the first two years of life. The results suggest that intensive household WASH and nutrition improvements alone may be insufficient to reduce *Shigella*/EIEC transmission and that complimentary interventions such as community level engineered systems that separate human excrement from the living environment or a vaccine would be required to meaningfully reduce transmission during the first two years of life in high transmission settings.

Current *Shigella* vaccines in development largely feature the O antigen of the lipopolysaccharide (LPS) as corresponding immunoglobin G (IgG) antibodies confer protection against shigellosis (17, 18) but more recent studies suggest the structural Ipa proteins, particularly IpaA, IpaB, IpaC and IpaD, may be correlates of protection (Ndungo et al., 2018), (Martinez-Becerra et al., 2012), (Nosrati et al., 2019), (Hashemi et al., 2024), (Bernshtein et al., 2022). Second-generation vaccines have begun to incorporate novel antigens, such as recombinant IpaB, IpaC and IpaD with LPS with high efficacy in mice (Mani et al., 2016), (Turbyfill et al., 2008), (Heine et al., 2013). Our results suggest that IpaA and IpaC may be important immunogens due to protein regions with high enrichment for linear epitopes. The limited enrichment from IpaB, a common target for vaccine design, relative to IpaA and IpaC may be because IpaB epitopes are largely conformational and therefore might not have been detected using the SERA PIWAS tiling method which is trained to detect linear peptides. Despite this, epitope motifs were detected in all Ipa proteins except IpaH3, many of which do not align with epitopes identified through traditional immunoinformatics in previous literature (Nosrati et al., 2019). These epitope motif families often corresponded to amino acid regions with high IgG enrichment, suggesting the SERA and IMUNE pipelines may be detecting novel epitopes not routinely detected through heuristic driven prediction methods. Validation of these epitopes through experimental methods are needed to confirm this.

In our setting, we estimated that maternal IgG anti-*Shigella*/EIEC Ipa proteins are present at high levels in early infancy and begins to decrease between ages 6 to 9-months. This suggests that childhood vaccination after 9-months may avoid immune blunting from maternal antibodies (Ndungo & Pasetti, 2020). This vaccine timing implied by IgG decay is similar to transplacental IgG seroprevalence against the lipopolysaccharide (LPS) antigen in a population of Zambian infants that suggested potential vaccination timing would be after 14 and before 52 weeks of age (Chisenga et al., 2021). It is also in agreement with a study based in Mali and Ethiopia that determined children in a 12-17 month age group were most vulnerable to *Shigella* infection using a multiplex immunoassay that included a panel of Ipa and LSP antigens. Our findings further support the current consensus that ideal vaccination window for maximizing protection is before 12-months in high burden settings. Vaccine crowding, in which multiple vaccines are given in a short amount of time, should be considered as this may diminish immune response to a *Shigella* vaccine (Bagamian et al., 2022).

This study had limitations. First, we were unable to characterize non-protein-based epitopes, such as the O-linked antigen of the lipopolysaccharide, due to the limitations of the protein based immunoinformatic pipeline. Though this is unlikely to have impacted our seroprevalence measure, it is possible we are underestimating true seropositivity by excluding IgG that is specific for non-Ipa *Shigella*/EIEC targets. Second, we did not investigate other isotypes of antibody responses, such as immunoglobin A (IgA). IgA is an important mucosal immunoglobin with similar levels to IgG during infection with *Shigella*/EIEC (De Alwis et al., 2021). It is possible there is additional epitope diversity from IgA that is not included in these results. Nevertheless, a key strength of this study is its randomized intervention assignment and longitudinal design, which allowed us to characterize the epitope repertoire within a community setting beyond laboratory-based animal models or computational prediction-based inference.

In summary, our findings suggest that WASH interventions alone were insufficient to reduce *Shigella*/EIEC transmission in rural Bangladesh based on IgG immune responses across Ipa proteins. Distinct age-specific antibody patterns over the first 28-months of life provide valuable insights that can inform vaccine development by identifying key motifs and epitopes relevant to the target population and estimating the rate of maternal IgG waning. Taken together, these results highlight the importance of timing for vaccine administration against *Shigella*/EIEC and suggest potential novel epitopes for vaccine design.

## Data Availability

All data needed to evaluate the conclusions in the paper are present in the paper and/or the Supplementary Materials. All data used to display the figures and the code used for analyses are available at (https://osf.io/u92pc).

## Acknowledgements

The authors thank Dr. Christine Tedijanto Wen for her contribution to the analysis plan.

## Author contributions

NW and BA conceived and designed the study. MZR, SA, AS, SF, SA, MSH, PM, MRD, AL, SL, DL collected and curated the data. MZ, KK, JR, and JS developed and performed the peptide display assay and analysis. NW and EK performed downstream computational and statistical analysis. RC assisted with data interpretation. NW wrote the first draft of the manuscript. All authors contributed to writing and approved the final manuscript.

## Supplemental Tables and Figures

**Figure S1:**
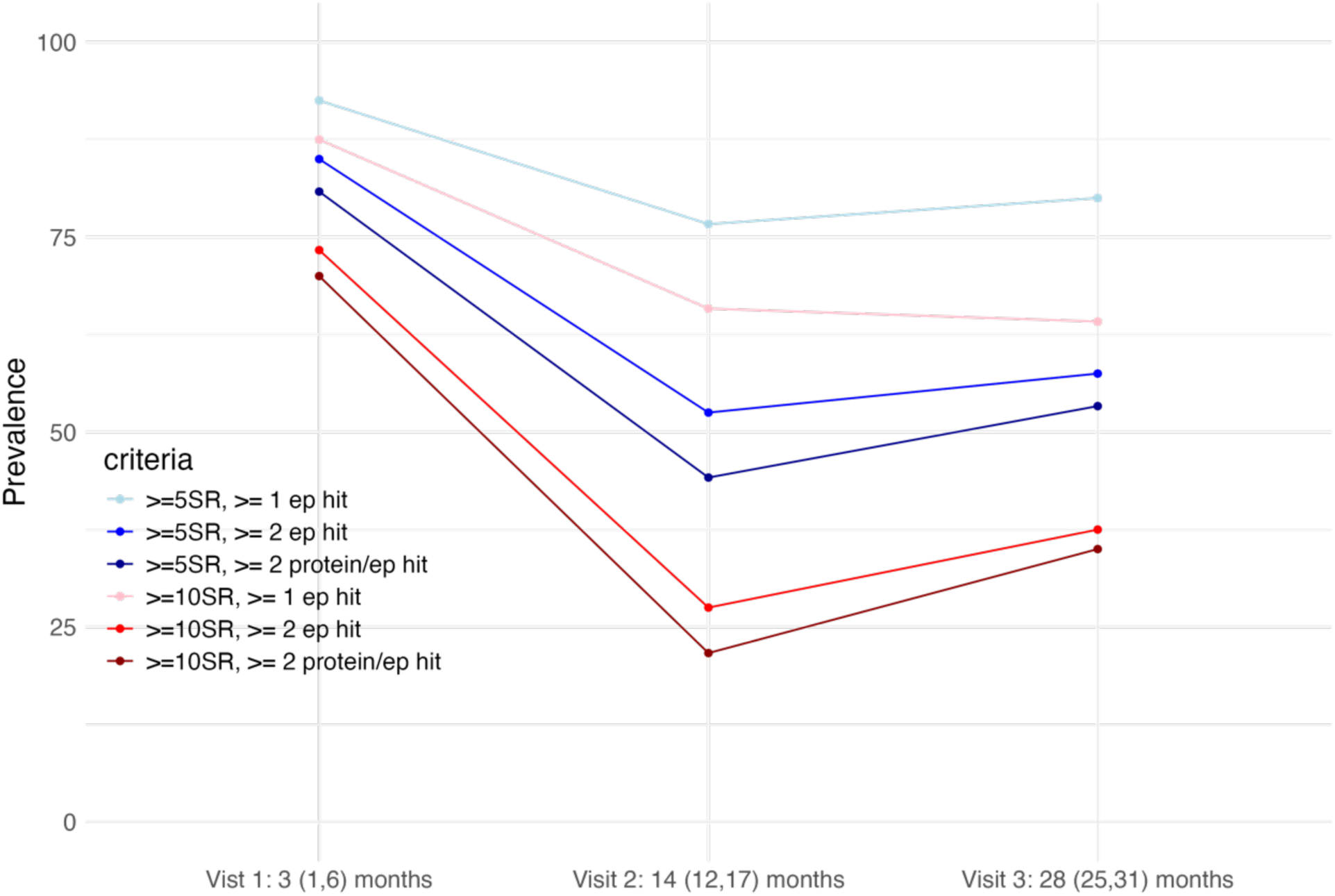
Sensitivity analysis of amino acid enrichment thresholds. Different thresholds for the assignment of seroreactive regions, defined by the number of individuals having an epitope hit in that region (at least five individuals or ten individuals), and the number of epitope hits within these assigned seroreactive regions to determine pathogen seropositivity (at least one epitope hit, at least two epitope hits, or at least two different proteins with one epitope hit each) were assessed. Seroprevalence across the three median ages: 3 months, 14 months and 28 months were compared. The final analysis defined seroreactive regions using a threshold of at least five individuals in the study sample having an epitope hit in the amino acid region, and at least two epitope hits in any seroreactive region across all Ipa proteins (navy line).

**Figure S2:**
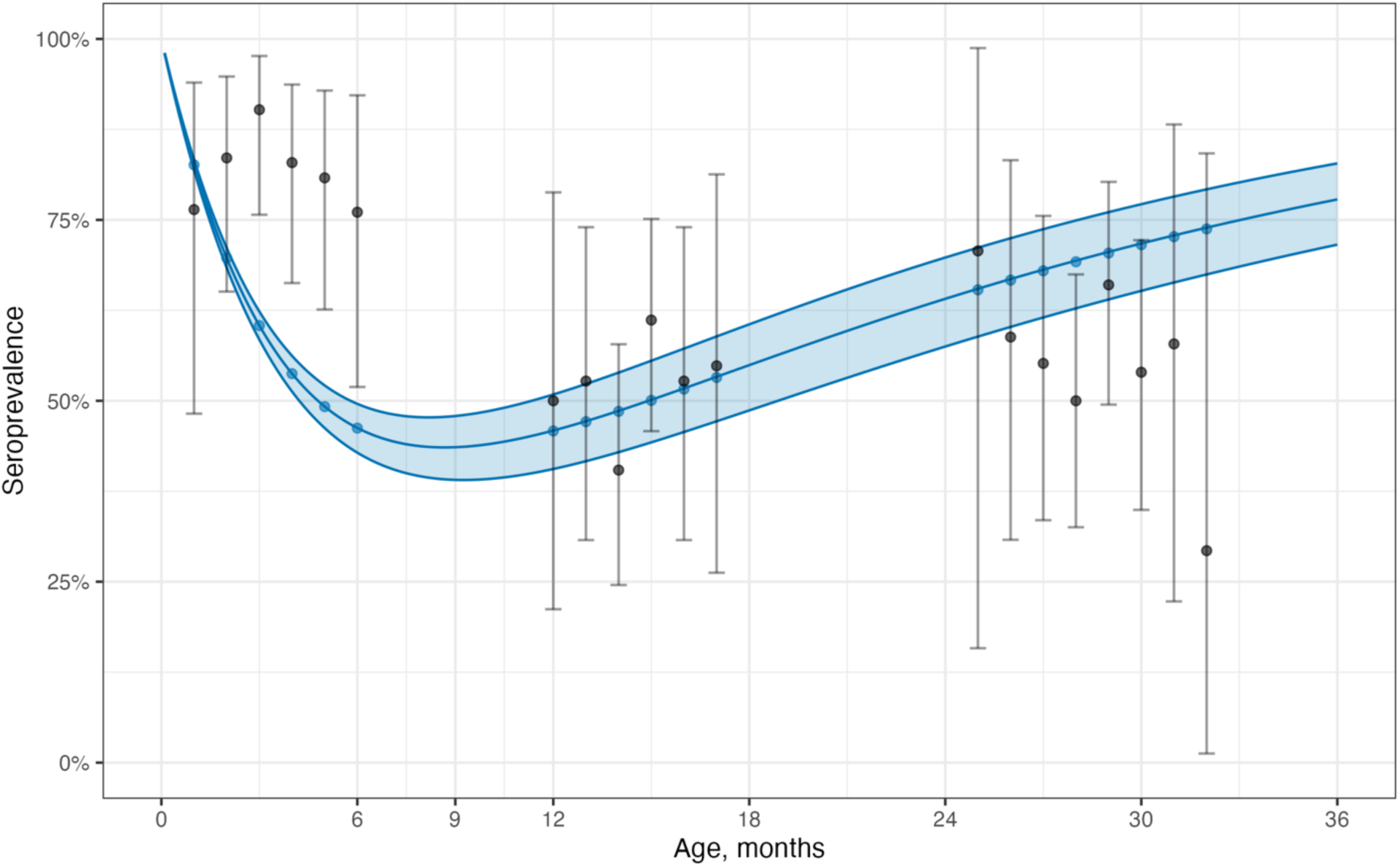
Model fit of IgG dynamics inferred from modified catalytic model. Black points represent the observed seroprevalence at each age in months from the 120 children in the cohort with 95% confidence interval. Blue points represent the estimated seroprevalence at each age in months, as inferred from the modified catalytic model. The blue line and ribbon represent the simulated model fit as well as the 95% confidence interval over 36 months.

---

**Table S1:**
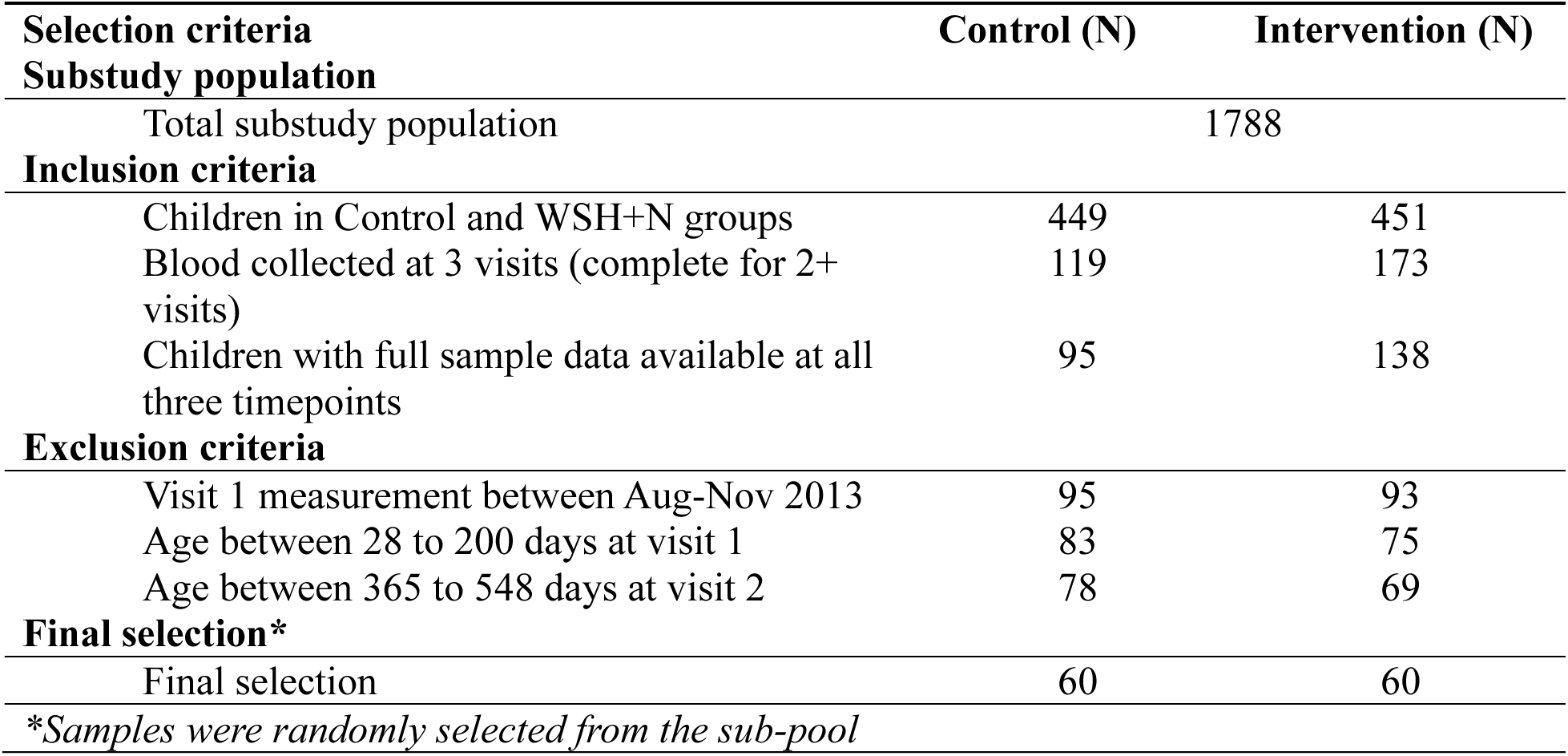
Random selection of study participants from the control and intervention groups. . Invention consisted of improved water, sanitation, handwashing conditions as well as improved nutrition (WSH+N).

**Table S2:** Shigella proteins and accession numbers used in panel.

| **Protein** | **Species** | **Accession number** |
| --- | --- | --- |
| IpaA | *Shigella sonnei* | Q3YTQ5 |
| IpaB | *Shigella sonnei* | Q3YTQ2 |
| IpaC | *Shigella sonnei* | Q3YTQ3 |
| IpaD | *Shigella sonnei* | Q3YTQ4 |
| IpaH3 | *Shigella sonnei* | Q3Z2I2 |

**Table S3.** Intervention effect: IgG Seroprevalence and seroconversion rate. Seroprevalence was defined as proportion of the population seropositive to *Shigella/*EIEC by having at least two epitope hits across all Ipa proteins (IpaA, IpaB, IpaC, IpaD, IpaH3). Seroconversion estimates based on 60 children in each group measured up to two times at median ages 14 and 28 months. Children who seroconverted by their second visit at median age 14 months were not included in the analysis of incident seroconversions between age 14 months and 28 months.

| Population measure | Control (95% CI) | Intervention (95% CI) | Measure of effect* |  | p-value |
| --- | --- | --- | --- | --- | --- |
|  |  |  | Difference (95% CI) | Ratio (95% CI) |  |
| Seroprevalence (%) | 56 (46, 67) | 53 (44, 62) | 3.0 (-10, 17) | 1.1 (0.8, 1.4) | 0.637 |
| Seroconversion rate (per 100 child-years) | 49.4 (39.4, 59.5) | 53.4 (42.4, 64.4) | 0.04 (-0.11, 0.19) | 1.1 (0.8, 1.4) | 0.601 |

\*Specific for population measure

**Table S4.** Intervention effect: IgG Quantitative measure values. Mean and max enrichment scores were calculated for amino acids within seroreactive regions, as previously defined in methods.

| Quantitative measure | Control (95% CI) | Intervention (95% CI) | Difference (95% CI) | p-value |
| --- | --- | --- | --- | --- |
| Mean IgG enrichment score | -0.16 (-0.19, -0.12) | -0.12 (-0.15, -0.10) | 0.03 (-0.0, 0.1) | 0.152 |
| Max IgG enrichment score | 3.94 (3.56, 4.32) | 3.83 (3.46, 4.20) | -0.11 (-0.6, 0.4) | 0.685 |
| IgG sera score | 35.04 (20.37, 49.71) | 35.47 (21.69, 49.25) | 0.43 (-19.7, 20.6) | 0.966 |

**Table S5.**
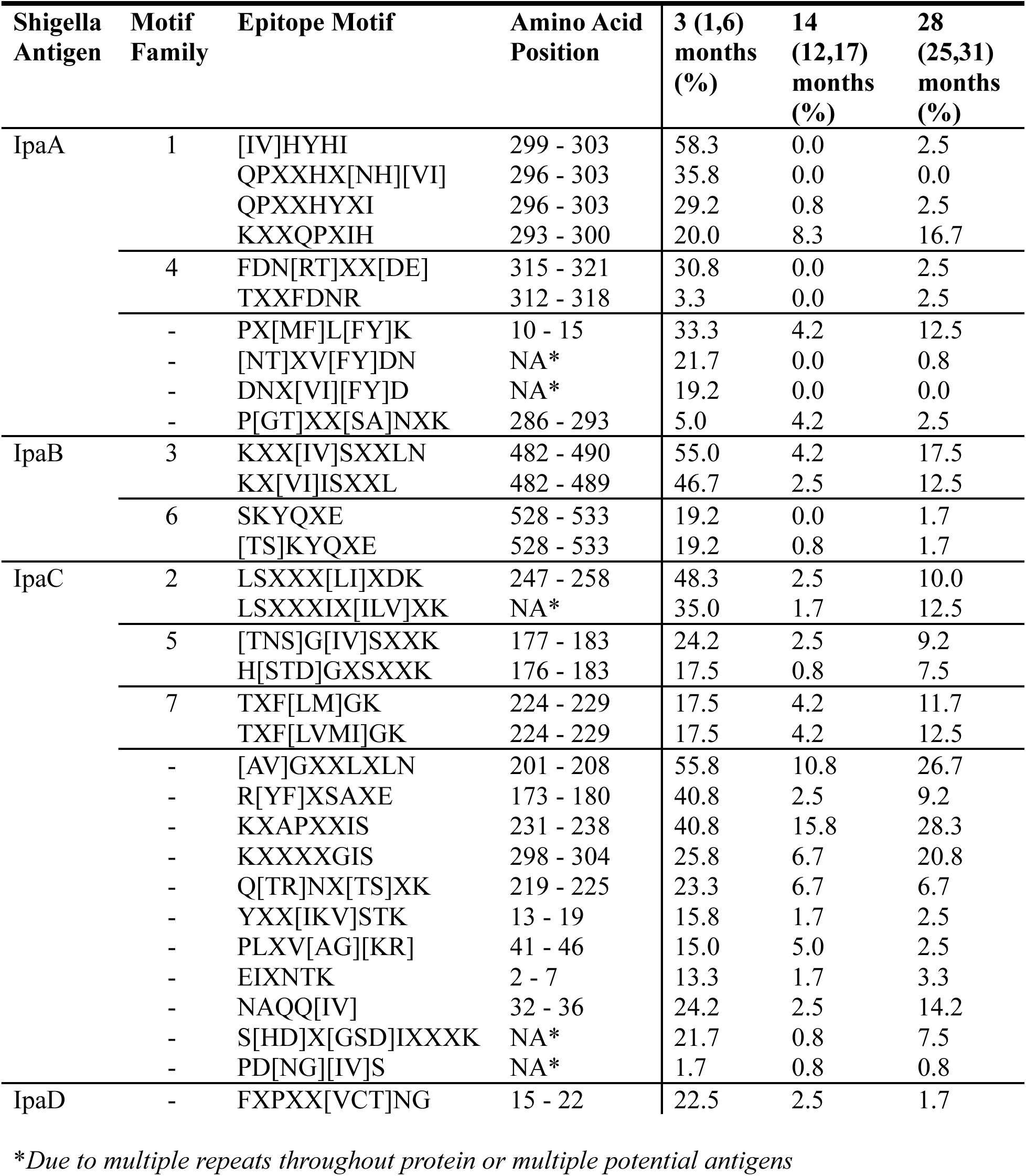
Epitope motifs detected across all Ipa proteins and proportion of samples positive to epitope, by visit. Motif families are numbered arbitrarily, according to the order they were discovered. Motif families only received a number if more than one epitope motif were detected in overlapping amino acid regions.

